# Relative effectiveness of BNT162b2, mRNA-1273, and Ad26.COV2.S vaccines and homologous boosting in preventing COVID-19 in adults in the US

**DOI:** 10.1101/2023.02.10.23285603

**Authors:** Van Hung Nguyen, Catherine Boileau, Alina Bogdanov, Meg Sredl, Mac Bonafede, Thierry Ducruet, Scott Chavers, Andrew Rosen, David Martin, Philip Buck, Diana Esposito, Nicolas Van de Velde, James A. Mansi

**Affiliations:** VHN Consulting Inc., Montreal, Quebec, Canada; Veradigm, Chicago, IL, USA; Moderna, Cambridge, MA, USA

**Keywords:** COVID-19, vaccine, BNT162b2, mRNA-1273, Ad26.COV2.S, booster, relative vaccine effectiveness

## Abstract

**Background:** Few head-to-head comparisons have been performed on the real-world effectiveness of COVID-19 booster vaccines. We evaluated the relative effectiveness (rVE) of a primary series of mRNA-1273 versus BNT162b2 and Ad26.COV2.S and a homologous mRNA booster against medically-attended, outpatient, and hospitalized COVID-19.

**Methods:** A dataset linking primary care electronic medical records with medical claims data was used for this retrospective cohort study of US patients ≥18 years vaccinated with a primary series between February and October 2021 (Part 1) and a homologous mRNA booster between October 2021 and January 2022 (Part 2). Adjusted hazard ratios (HR) were derived from 1:1 matching adjusted across potential covariates. rVE was (1-HR_adjusted_) x 100. Additional analysis was performed across regions and age groups.

**Results:** Following adjustment, Part 1 rVE for mRNA-1273 versus BNT162b2 was 23% (95% CI: 22%–25%), 23% (22%–25%), and 19% (14%–24%) whilst the rVE for mRNA-1273 versus Ad26.COV2.S was 50% (48%–51%), 50% (48%–52%), and 57% (53%–61%) against any medically-attended, outpatient, and hospitalized COVID-19, respectively. The adjusted rVE in Part 2 for mRNA-1273 versus BNT162b2 was 14% (10%–18%), 13% (8%– 17%), and 19% (1%–34%) against any medically-attended, outpatient, and hospitalized COVID-19, respectively. rVE against medically-attended COVID-19 was higher in adults ≥65 years (35%; 24%–47%) than those 18–64 years (13%; 9%–17%) after the booster.

**Conclusions:** In this study, mRNA-1273 was more effective than BNT162b2 or Ad26.COV2.S following primary series during a Delta-dominant period, and than BNT162b2 as a booster during an Omicron-dominant period.

**Key points:** mRNA-1273 was associated with a lower risk than BNT162b2 or Ad26.COV2.S of any medically-attended, outpatient, or hospitalized COVID-19 after primary series and of any medically-attended, outpatient, or hospitalized COVID-19 vs BNT162b2 after a homologous mRNA booster

## Introduction

During the initial wave of COVID-19 vaccinations in late 2020 and early 2021, three SARS-CoV-2 vaccines were made available in the US under emergency use authorizations (EUA): two mRNA vaccines (mRNA-1273 and BNT162b2, available as two-dose primary series and boosters) and an adenovirus-based vaccine (Ad26.COV2.S; available as a one-dose primary series and booster). Data from clinical trials indicated high vaccine efficacy through to 6 months post-vaccination, particularly for mRNA vaccines [1-7]. Following the large-scale roll-out of these vaccines to the US population, COVID-19-related hospitalizations and deaths decreased rapidly in the first half of 2021; however, the emergence of the Delta variant followed by the Omicron variant and sub-variants resulted in a resurgence of cases [8]. Viral mutations rendering these variants more fit to evade the immune response and increased transmission, combined with host factors associated with greater risk for COVID-19-related morbidity and mortality, contributed to the observed waning of vaccine effectiveness [9-12]. However, administration of an mRNA vaccine booster dose was shown to increase effectiveness against symptomatic and severe disease during both the periods of Delta and Omicron predominance [13]. From October 2021 onwards, mRNA booster doses have been recommended for at-risk populations and subsequently expanded to all individuals over 12 years of age who have completed a primary COVID-19 vaccine series [14, 15].

Immunisation with a primary series followed by boosting with mRNA-1273 and BNT162b2 in adults ≥18 years, allows for a direct comparison of vaccine effectiveness. Such data will be important to support vaccination decision making, particularly in settings where vaccine effectiveness is a criterion for selection.

A limited number of studies have specifically compared the effectiveness of the two mRNA vaccines during periods when Delta and Omicron variants predominated [16-18]. However, no study to date has evaluated vaccine effectiveness within the same cohort from primary vaccination through to booster spanning both Delta- and Omicron-dominant periods. While some comparative effectiveness studies have been published, direct head-to-head effectiveness research is needed particularly within the context of emerging variants and updated formulations. To address this, we conducted a retrospective longitudinal study using a large integrated electronic health record (EHR) [19] dataset to assess the relative vaccine effectiveness (rVE) of mRNA-1273 following primary and booster vaccination versus BNT162b2 and Ad26.COV2.S in preventing COVID-19-related medical encounters (outpatient and hospitalized COVID-19) during periods where Delta and Omicron variants predominated in the United States.

## Methods

### Study design

This retrospective observational longitudinal study was conducted between February 2021 and January 2022 using de-identified (see supplement for details) electronic medical records from primary care and specialist clinics with linked pharmacy and medical claims data. Data were evaluated for adults aged ≥18 years with a record of receiving a full primary series of mRNA-1273, BNT162b2, or Ad26.COV2.S between 1st February 2021 and 18th October 2021 (Part 1) and a homologous mRNA booster (mRNA-1273 and BNT162b2 only) between 19^th^ October 2021 and 31^st^ January 2022 (Part 2; **Figure 1**). A cut-off date between the primary series and booster vaccination of 19^th^ October 2021 was based on the date on which CDC recommendations for booster doses came into effect. Only individuals who were included in Part 1 were eligible for inclusion in Part 2 of the study. The study was designed, implemented, and reported in accordance with Good Pharmacoepidemiological Practices, applicable local regulations, and the ethical principles laid down in the Declaration of Helsinki. Study findings are reported in accordance with the Reporting of Studies Conducted Using Observational Routinely Collected Health Data recommendations.

**Figure 1.**
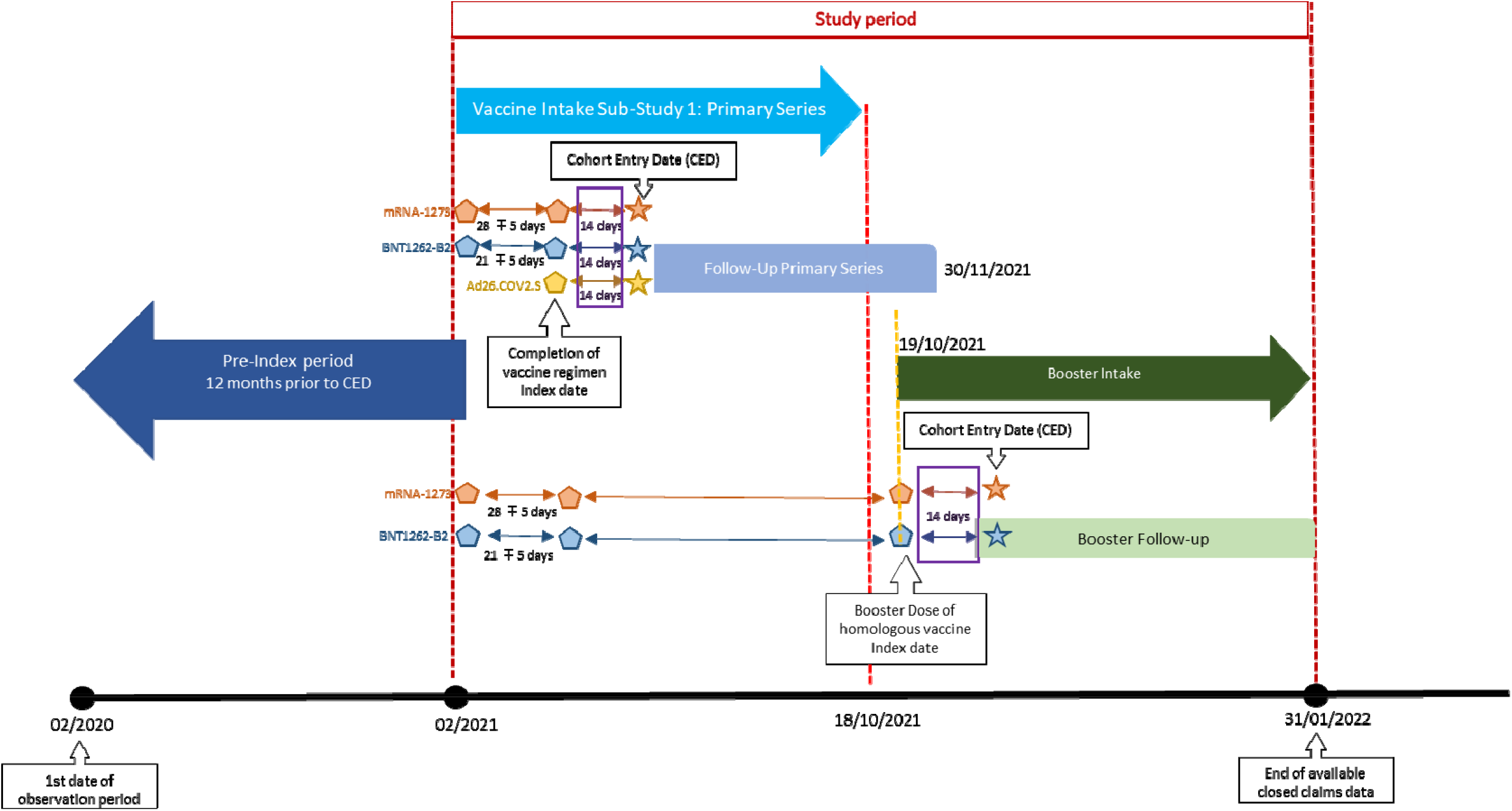
Study design

### Data source

This analysis was performed on an integrated real-world EHR dataset (Veradigm Health Insights) together with pharmacy and medical claims data (Komodo Health Inc., San Francisco, CA, USA). This integrated dataset has been used extensively to evaluate vaccine effectiveness [20-22] and has been shown to be representative of the US population containing key variables for conducting RWE research [19]. The Veradigm Health Insights EHR database contains data on healthcare interactions for over 55 million patients in the US whose providers use the Allscripts Touchworks, Allscripts PRO, and Practice Fusion EHRs, including details of prescriptions and vaccinations for both primary care physicians and specialists. Closed medical claims data were included, i.e., adjudicated claims within the period in which the patient was continuously enrolled in an insurance plan. As a noninterventional, retrospective database study using a certified Health Insurance Portability and Accountability Act–compliant deidentified research database, approval by an institutional review board was not necessary.

### Study population

Individuals were eligible for inclusion in Part 1 of the study if they were ≥18 years of age, had received two doses of mRNA-1273 or BNT162b2, or one dose of Ad26.COV2.S (using CPT codes; **Supplementary Table 1**), had linked EHR and claims data together with EHR activity >365 days before the index date (**Figure 1**). The index date (14 days prior to the cohort entry date) was defined as the date of receipt of the second mRNA vaccine dose, or the date of receipt of the single Ad26.COV2.S dose. Receipt of a homologous COVID-19 vaccine booster (see **Supplementary Table 1**) between 19^th^ October and 31^st^ January 2022 was an additional criterion for Part 2 of the study; individuals who received a booster dose prior to 19^th^ October 2021 were excluded. Exclusion criteria included receipt of a heterologous COVID-19 vaccine, no record of the second dose of mRNA-1273 within 28±5 days of the first dose or BNT162b2 within 21±5 days of the first dose, evidence of previous confirmed COVID-19 infection between 1^st^ January 2020 and the beginning of the follow-up period, homologous vaccination between the date of complete vaccine regimen and full vaccination date (i.e. 14 days following the completed vaccine regimen), and missing birth year or gender.

Fully vaccinated mRNA-1273 recipients, i.e., those who had completed the primary series, were matched 1:1 with individuals from the comparator vaccine groups (BNT162b2 and AD26.COV2.S for Part 1; BNT162b2 only for Part 2) based on sex, geographic region, and race. Age was matched within five-year age groups and date of full vaccination was matched ±5 days. mRNA-1273 recipients could be included in both primary series comparisons (BNT162b2 and Ad26.COV2.S).

### Exposure and outcome ascertainment

EHR data, together with pharmacy and medical claims data, were used to identify individuals vaccinated between the cohort entry and end of intake dates (**Figure 1**). Follow-up for assessment of outcomes of interest was performed from 14 days after the index date (defined as “full vaccination”) until the earliest of the following events: first occurrence of an outcome of interest; end of the observation period; or disenrollment from their medical/pharmacy plan. Cohort entry date for Part 2 was the date of receipt of the booster dose and the follow-up period ranged from the cohort entry date to 31^st^ January 2022.

The primary outcome was any medically-attended COVID-19, defined as any medical encounter with a COVID-19 diagnosis. Secondary outcomes were hospitalized and outpatient COVID-19 (see **Supplementary Table 2** for defining codes).

### Statistical analysis

Covariate balance at baseline for each comparator vaccine versus mRNA-1273 was assessed using standardized mean differences (SMDs) prior to any adjustments. Kaplan Meier plots with associated 95% confidence intervals were generated to assess rVE over time. Right censoring was defined as the end of the follow-up period, with a maximum follow-up time of 265 days post-primary series and 90 days post-booster. For each of the comparisons, unadjusted HRs were estimated using a univariate Cox regression model with no covariates. Adjusted HRs were calculated using a multivariable Cox regression model, adjusting for matching variables and covariates with an SMD ≥0.1 (**Supplementary Tables 3 and 4**). rVE was defined as (1-adjusted HR) x 100. Statistical analyses were performed using R Statistical Software (v4.1.3) [23] and the *survival* (v3.2-13; Therneau, 2021) and *MatchIt* (v4.3.2; Ho et al., 2021) packages.

### Additional exploratory analyses

Exploratory analysis was also performed for the primary and secondary outcomes of interest by age group (18–64 years, 65–74 years, ≥65 years, and ≥75 years) and region (Midwest, Northeast, South, West, or unknown). A sensitivity analysis was also performed for laboratory-confirmed SARS-CoV-2 infection for Parts 1 and 2, defined as a positive lab result as recorded in the EHR.

## Results

Of the ∼15.9 million patients included in the linked EHR-claims dataset who had been fully vaccinated with a primary series BNT162b2, mRNA-1273, or Ad26.COV2.S between 1st February and 18th October 2021, 4,404,091 were included in the first part of this analysis (**Figure 2a**). Among these, 2,092,304 received BNT162b2, 1,788,220 received mRNA-1273, and 523,567 received Ad26.COV2.S. Overall, 1,529,930 individuals who received mRNA-1273 were matched with BNT162b2 recipients, and 484,795 were matched with Ad26.COV2.S recipients (**Table 1; Supplementary Table 3**).

**Figure 2.**
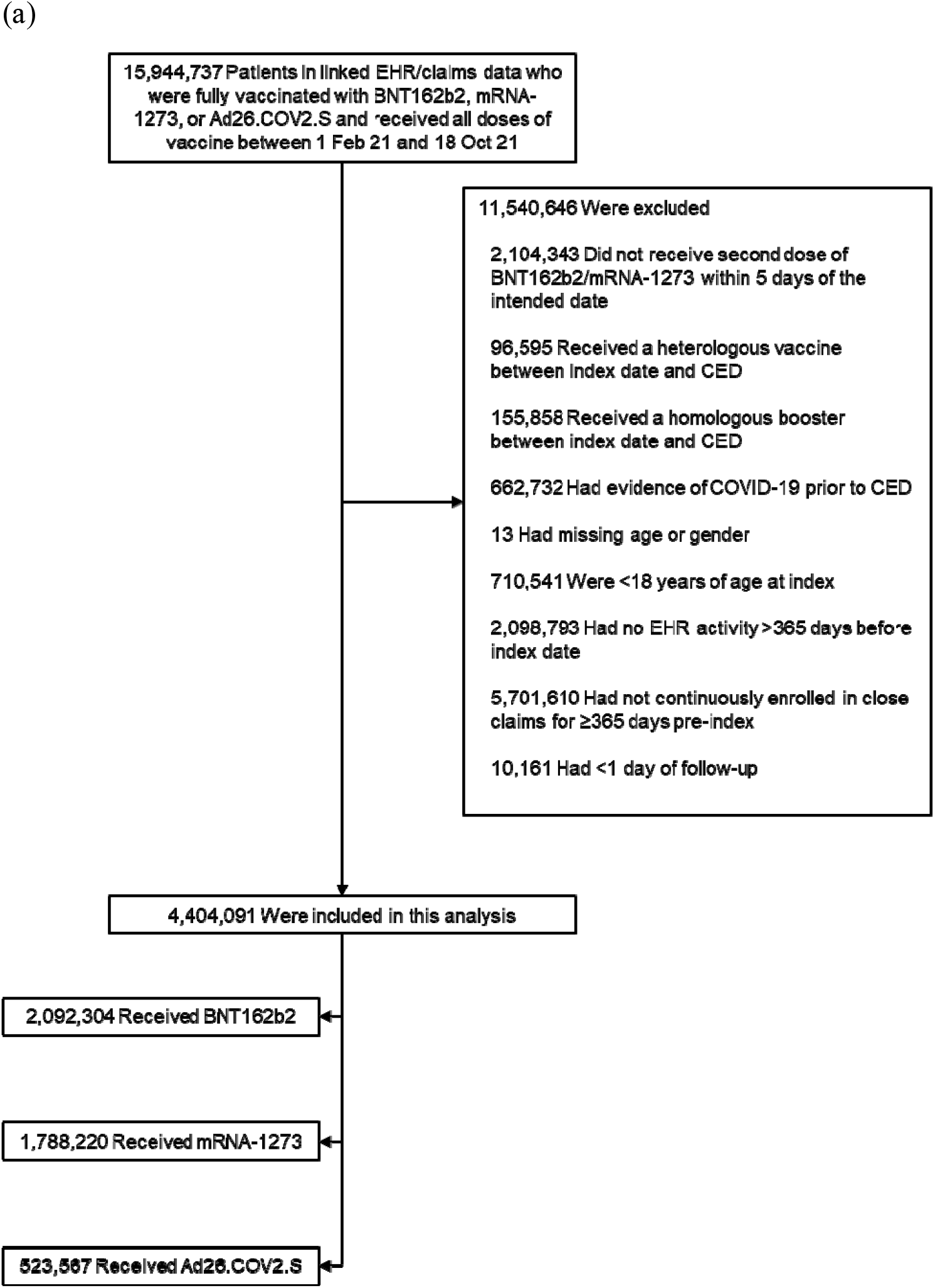
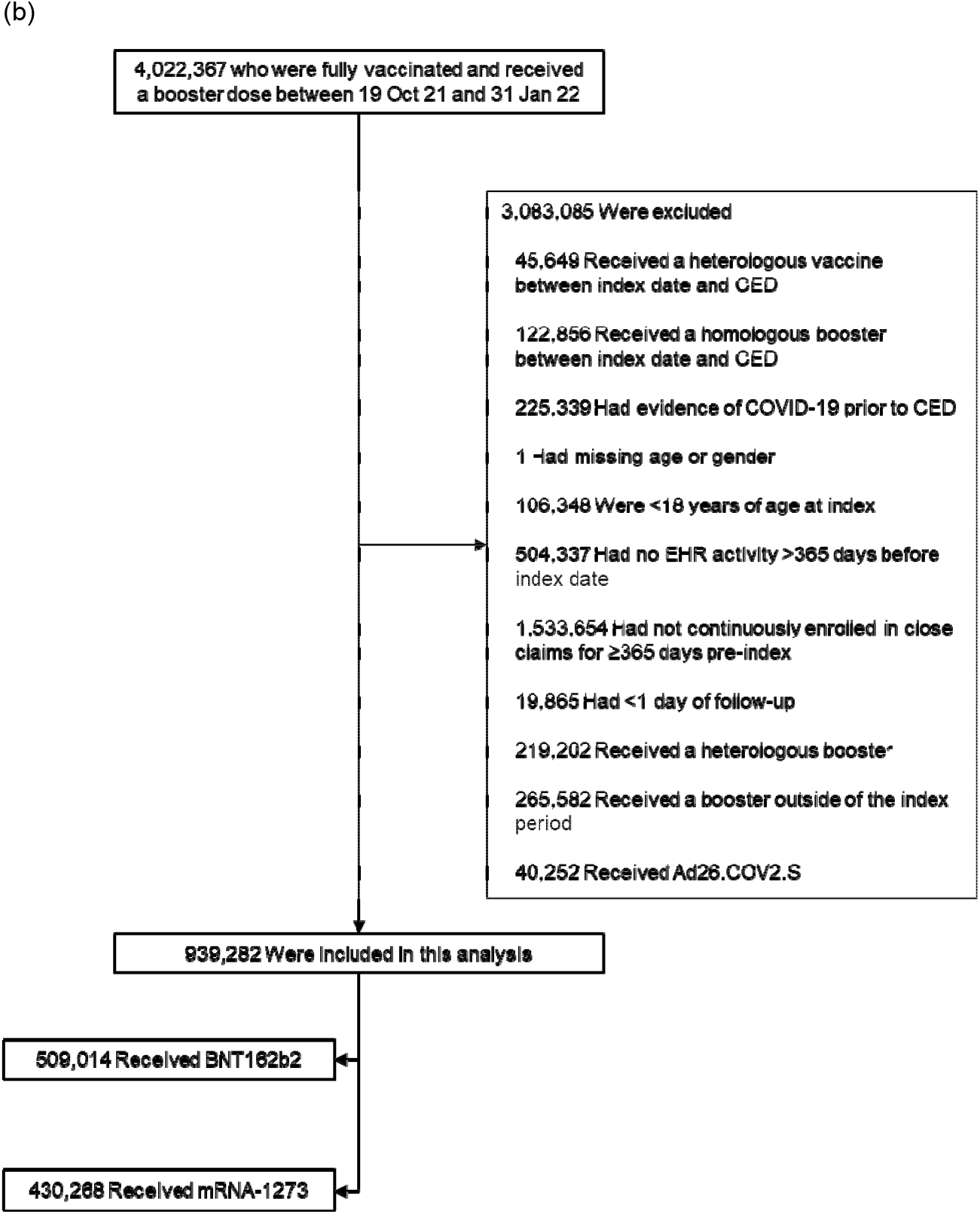
Selection of (a) BNT162b2, mRNA-1273, and Ad26.COV2.S primary series recipients and (b) BNT162b2 and mRNA-1273 booster recipients for inclusion in the analysis CED, cohort entry date; EHR, electronic health record Note: exclusion criteria were evaluated in a step-wise fashion, summing to the total excluded

**Table 1.**
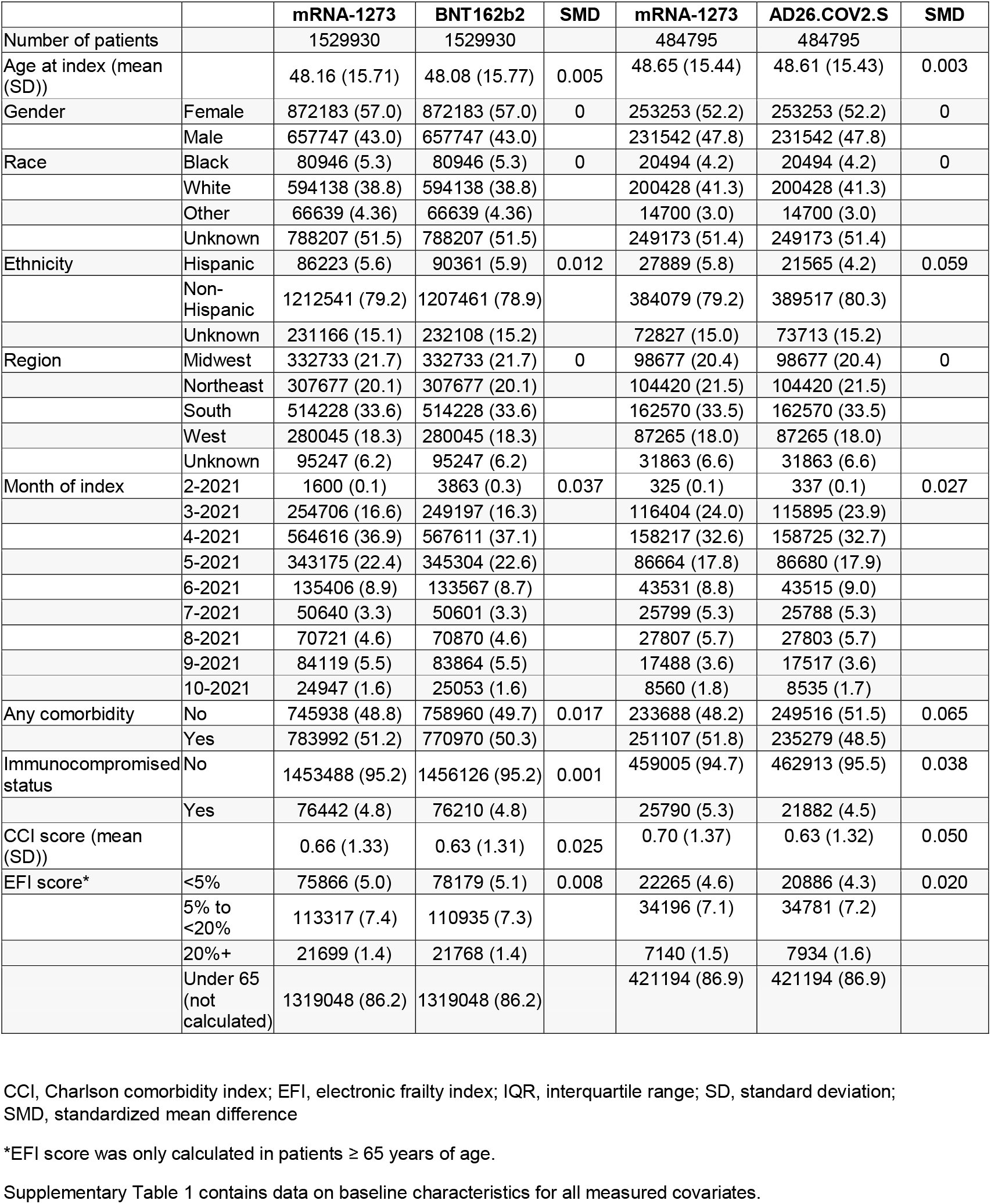
Key baseline characteristics of matched patients included in the comparisons of mRNA-1273 vs BNT162b2 and Ad26.COV2.S after the primary series

In total, 4,022,367 individuals received a booster dose of BNT162b2 or mRNA-1273 between 19th October 2021 and 31st January 2022 (**Figure 2b**). Of these, 509,014 BNT162b2 and 430,268 mRNA-1273 recipients were eligible for inclusion. Of these, 368,100 matched individuals who received a homologous booster dose of mRNA.1273 or BNT162b2 were included in the second part of the analysis (**Table 2; Supplementary Table 4**).

**Table 2.**
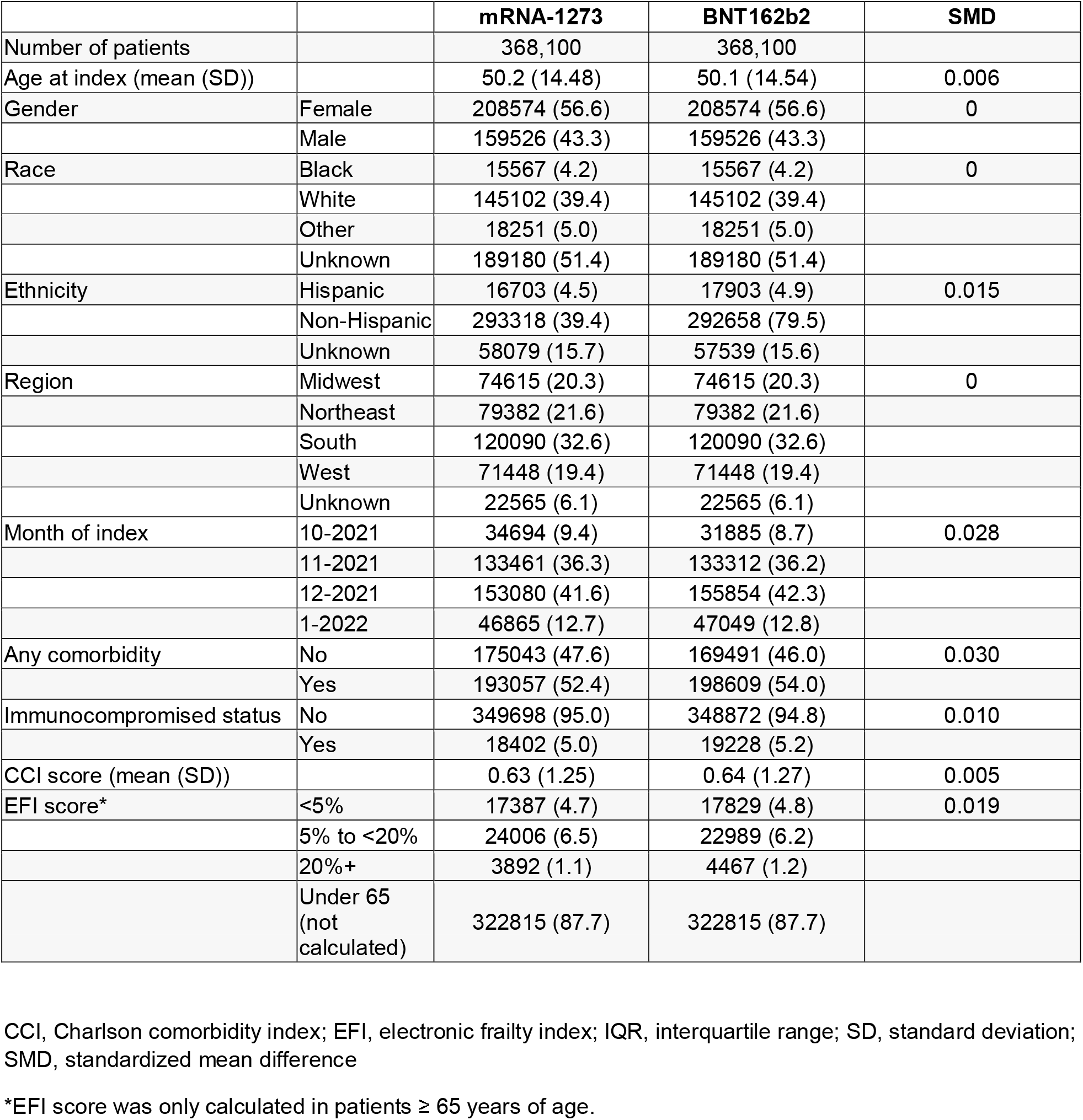
Key baseline characteristics of matched patients included in the comparison of mRNA-1273 with BNT162b2 after a booster dose

### Cumulative incidence of COVID-19

Analysis of the primary outcome of any medically-attended COVID-19 showed a lower cumulative incidence for mRNA-1273 compared with BNT162b2 or Ad26.COV2.S after the primary series, with adjusted HRs of 0.77 (95% CI: 0.75–0.78) and 0.50 (0.49–0.52), respectively (**Supplementary Figure 1, Table 3**). This corresponded to rVE estimates of 23% (22%–25%) and 50% (48%–51%), respectively.

**Table 3.** Unadjusted and adjusted hazard ratios for mRNA-1273 with BNT162b2 and Ad26.COV2.S (primary series) and with BNT162b2 (booster dose)

|  | Primary Series |  |  |  | Booster |  |
| --- | --- | --- | --- | --- | --- | --- |
|  | mRNA-1273 vs BNT162b2 |  | mRNA-1273 vs Ad26.COV2.S |  | mRNA-1273 vs BNT162b2 |  |
| Outcome type | Unadjusted HR | Adjusted HR | Unadjusted HR | Adjusted HR | Unadjusted HR | Adjusted HR |
| Any medically-attended COVID-19<sup>a</sup> | 0.76 (0.75 - 0.78) | 0.77 (0.75 - 0.78) | 0.50 (0.49 - 0.52) | 0.50 (0.49 - 0.52) | 0.81 (0.78 – 0.85) | 0.86 (0.82 – 0.90) |
| Outpatient COVID-19<sup>b</sup> | 0.76 (0.74 - 0.78) | 0.77 (0.75 - 0.78) | 0.50 (0.48 - 0.52) | 0.50 (0.48 - 0.52) | 0.83 (0.79 – 0.87) | 0.87 (0.83 – 0.92) |
| COVID-19 Hospitalization<sup>c</sup> | 0.84 (0.79 - 0.90) | 0.81 (0.76 - 0.86) | 0.43 (0.39 - 0.47) | 0.43 (0.39 - 0.47) | 0.77 (0.63 – 0.94) | 0.81 (0.66 – 0.99) |

<sup>a</sup>Defined as any medical encounter with a COVID-19 diagnosis or positive COVID-19 laboratory test
<sup>b</sup>Defined as a hospitalization where COVID-19 was listed in any diagnosis position
<sup>c</sup>Defined as an encounter recorded either in the EHR or on a claim that is not a hospitalization claim
Details of the codes used to identify COVID-19-related medical encounters are provided in Supplementary Table 2

For both vaccine comparisons, adjusted HRs across the secondary outcomes analysed reflected those for medically-attended COVID-19 (**Table 3**). For mRNA-1273 versus BNT162b2, adjusted rVE ranged from 23%–28% for hospitalized and outpatient COVID-19, and for mRNA-1273 versus Ad26.COV2.S adjusted rVE was 50% for both measures.

Analysis of mRNA-1273 versus BNT162b2 after the booster dose resulted in an rVE of 14% (10%–18%) for the primary outcome of any medically attended COVID-19 (**Table 3, Supplementary Figure 2**). Estimates of adjusted HR for outpatient and hospitalized COVID-19 also showed a higher VE for mRNA-1273 compared with BNT162b2, with an rVE of 13% (8%–17%) and 19% (1%–34%), respectively.

### Analysis by age group

Overall, 75.8–76.8% of individuals were in the 18–64 years age group across comparisons of the primary series, and 87.7% for the booster dose (**Supplementary Table 5**).

In the exploratory analysis by age, after the primary series, adjusted HRs for mRNA-1273 versus BNT162b2 were highest for the primary and secondary outcomes in the 18–64 years age group (0.78–0.88), although confidence intervals overlapped between all age groups (18– 64, 65–74, and ≥75), suggesting no clear age effect (**Table 4**). Similarly, the data suggested no clear age effect for mRNA-1273 versus Ad26.COV2.S, with adjusted HRs across outcomes and age groups. Across all outcomes and for all age groups, higher point estimates for rVE of mRNA-1273 was observed as compared to the other two vaccines (rVE range: 12%–67%).

**Table 4.** Unadjusted and adjusted hazard ratios (95% confidence interval) by age group for the any COVID-19 and outpatient, hospitalized, and lab-confirmed COVID-19 for the comparisons of mRNA-1273 with BNT162b2 and Ad26.COV2.S (primary series) and with BNT162b2 (booster dose)

|  | Primary Series |  |  |  | Booster |  |
| --- | --- | --- | --- | --- | --- | --- |
|  | mRNA-1273 vs BNT162b2 |  | mRNA-1273 vs Ad26.COV2.S |  | mRNA-1273 vs BNT162b2 |  |
| Outcome | Unadjusted | Adjusted | Unadjusted | Adjusted | Unadjusted | Adjusted |
| **18-64 years** |  |  |  |  |  |  |
| Overall | 0.78 (0.76 - 0.80) | 0.78 (0.76 - 0.80) | 0.51 (0.49 - 0.53) | 0.51 (0.49 - 0.53) | 0.85 (0.80 – 0.89) | 0.87 (0.83 – 0.91) |
| Outpatient | 0.77 (0.75 - 0.79) | 0.78 (0.76 - 0.79) | 0.51 (0.49 - 0.53) | 0.51 (0.49 - 0.53) | 0.86 (0.82 – 0.91) | 0.89 (0.84 – 0.94) |
| Hospitalizations | 0.91 (0.84 - 0.99) | 0.88 (0.81 - 0.95) | 0.44 (0.39 - 0.50) | 0.44 (0.39 - 0.50) | 0.86 (0.68 – 1.09) | 0.88 (0.70 – 1.12) |
| **≥65 years** |  |  |  |  |  |  |
| Overall | 0.70 (0.66 - 0.73) | 0.70 (0.66 - 0.74) | 0.47 (0.43 - 0.51) | 0.47 (0.43 - 0.51) | 0.60 (0.52 – 0.93) | 0.65 (0.56 – 0.76) |
| Outpatient | 0.69 (0.65 - 0.73) | 0.70 (0.66 - 0.74) | 0.46 (0.42 - 0.51) | 0.46 (0.42 - 0.51) | 0.61 (0.52 – 0.71) | 0.66 (0.57 – 0.78) |
| Hospitalizations | 0.71 (0.64 - 0.80) | 0.71 (0.64 - 0.80) | 0.41 (0.34 - 0.48) | 0.41 (0.34 - 0.48) | 0.57 (0.38 – 0.83) | 0.60 (0.40 – 0.89) |
| **65-74 years** |  |  |  |  |  |  |
| Overall | 0.70 (0.65 - 0.74) | 0.70 (0.66 - 0.75) | 0.49 (0.44 - 0.55) | 0.49 (0.44 - 0.55) | 0.59 (0.49 – 0.70) | 0.64 (0.53 – 0.76) |
| Outpatient | 0.69 (0.64 - 0.74) | 0.70 (0.65 - 0.75) | 0.47 (0.42 - 0.52) | 0.47 (0.42 - 0.52) | 0.59 (0.49 – 0.71) | 0.64 (0.53 – 0.77) |
| Hospitalizations | 0.80 (0.69 - 0.93) | 0.79 (0.68 - 0.92) | 0.46 (0.37 - 0.57) | 0.46 (0.37 - 0.57) | 0.54 (0.33 – 0.89) | 0.55 (0.33 – 0.91) |
| **≥75 years** |  |  |  |  |  |  |
| Overall | 0.69 (0.63 - 0.76) | 0.70 (0.64 - 0.77) | 0.42 (0.36 - 0.49) | 0.42 (0.36 - 0.49) | 0.63 (0.48 – 0.84) | 0.73 (0.55 – 0.97) |
| Outpatient | 0.70 (0.63 - 0.78) | 0.71 (0.64 - 0.79) | 0.44 (0.37 - 0.52) | 0.44 (0.37 - 0.52) | 0.67 (0.50 – 0.90) | 0.75 (0.55 – 1.01) |
| Hospitalizations | 0.62 (0.52 - 0.73) | 0.63 (0.54 - 0.75) | 0.33 (0.25 - 0.44) | 0.33 (0.25 - 0.44) | 0.62 (0.34 – 1.13) | 0.69 (0.37 - 1.28) |

NA, not assessable

After the booster dose, a trend for an age-effect was observed against any medically-attended and outpatient COVID-19, with lower HRs in individuals ≥65 years of age compared with younger adults. Point estimates were also lower for the other outcomes analyzed across age groups; however, confidence intervals overlapped (**Table 4**).

### Analysis by region

In the exploratory analysis by regin, no apparent differences in adjusted HRs were evident when assessed by region for either the primary series or booster dose (**Table 5**). For mRNA-1273 versus BNT162b2, rVE for the primary outcome of any medically-attended COVID-19 after the primary series ranged from 20%–24% across regions, with overlapping confidence intervals between regions for secondary outcomes measures. Results for the mRNA-1273 versus Ad26.COV2.S comparison also appeared similar across regions, with rVE for any medically-attended COVID-19 ranging from 46%–52%. After the homologous booster, rVE for mRNA-1273 versus BNT162b2 ranged from 4%–29% across the three measures, with overlapping confidence intervals between regions.

**Table 5.**
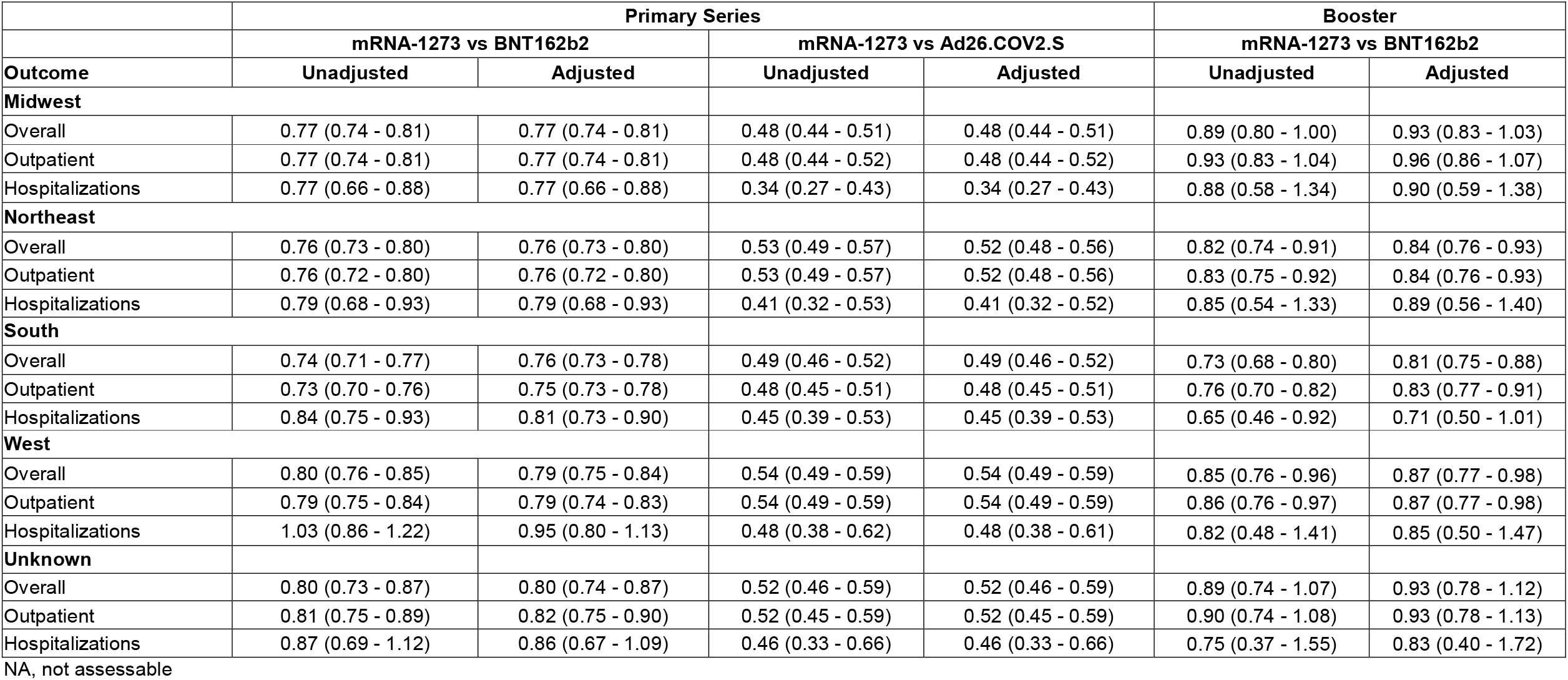
Unadjusted and adjusted hazard ratios (95% confidence interval) by region for the any COVID-19 and outpatient, hospitalized, and lab-confirmed COVID-19 for the comparisons of mRNA-1273 with BNT162b2 and Ad26.COV2.S (primary series) and with BNT162b2 (booster dose)

|  | Primary Series |  |  |  | Booster |  |
| --- | --- | --- | --- | --- | --- | --- |
|  | mRNA-1273 vs BNT162b2 |  | mRNA-1273 vs Ad26.COV2.S |  | mRNA-1273 vs BNT162b2 |  |
| Outcome | Unadjusted | Adjusted | Unadjusted | Adjusted | Unadjusted | Adjusted |
| **Midwest** |  |  |  |  |  |  |
| Overall | 0.77 (0.74 - 0.81) | 0.77 (0.74 - 0.81) | 0.48 (0.44 - 0.51) | 0.48 (0.44 - 0.51) | 0.89 (0.80 - 1.00) | 0.93 (0.83 - 1.03) |
| Outpatient | 0.77 (0.74 - 0.81) | 0.77 (0.74 - 0.81) | 0.48 (0.44 - 0.52) | 0.48 (0.44 - 0.52) | 0.93 (0.83 - 1.04) | 0.96 (0.86 - 1.07) |
| Hospitalizations | 0.77 (0.66 - 0.88) | 0.77 (0.66 - 0.88) | 0.34 (0.27 - 0.43) | 0.34 (0.27 - 0.43) | 0.88 (0.58 - 1.34) | 0.90 (0.59 - 1.38) |
| **Northeast** |  |  |  |  |  |  |
| Overall | 0.76 (0.73 - 0.80) | 0.76 (0.73 - 0.80) | 0.53 (0.49 - 0.57) | 0.52 (0.48 - 0.56) | 0.82 (0.74 - 0.91) | 0.84 (0.76 - 0.93) |
| Outpatient | 0.76 (0.72 - 0.80) | 0.76 (0.72 - 0.80) | 0.53 (0.49 - 0.57) | 0.52 (0.48 - 0.56) | 0.83 (0.75 - 0.92) | 0.84 (0.76 - 0.93) |
| Hospitalizations | 0.79 (0.68 - 0.93) | 0.79 (0.68 - 0.93) | 0.41 (0.32 - 0.53) | 0.41 (0.32 - 0.52) | 0.85 (0.54 - 1.33) | 0.89 (0.56 - 1.40) |
| **South** |  |  |  |  |  |  |
| Overall | 0.74 (0.71 - 0.77) | 0.76 (0.73 - 0.78) | 0.49 (0.46 - 0.52) | 0.49 (0.46 - 0.52) | 0.73 (0.68 - 0.80) | 0.81 (0.75 - 0.88) |
| Outpatient | 0.73 (0.70 - 0.76) | 0.75 (0.73 - 0.78) | 0.48 (0.45 - 0.51) | 0.48 (0.45 - 0.51) | 0.76 (0.70 - 0.82) | 0.83 (0.77 - 0.91) |
| Hospitalizations | 0.84 (0.75 - 0.93) | 0.81 (0.73 - 0.90) | 0.45 (0.39 - 0.53) | 0.45 (0.39 - 0.53) | 0.65 (0.46 - 0.92) | 0.71 (0.50 - 1.01) |
| **West** |  |  |  |  |  |  |
| Overall | 0.80 (0.76 - 0.85) | 0.79 (0.75 - 0.84) | 0.54 (0.49 - 0.59) | 0.54 (0.49 - 0.59) | 0.85 (0.76 - 0.96) | 0.87 (0.77 - 0.98) |
| Outpatient | 0.79 (0.75 - 0.84) | 0.79 (0.74 - 0.83) | 0.54 (0.49 - 0.59) | 0.54 (0.49 - 0.59) | 0.86 (0.76 - 0.97) | 0.87 (0.77 - 0.98) |
| Hospitalizations | 1.03 (0.86 - 1.22) | 0.95 (0.80 - 1.13) | 0.48 (0.38 - 0.62) | 0.48 (0.38 - 0.61) | 0.82 (0.48 - 1.41) | 0.85 (0.50 - 1.47) |
| **Unknown** |  |  |  |  |  |  |
| Overall | 0.80 (0.73 - 0.87) | 0.80 (0.74 - 0.87) | 0.52 (0.46 - 0.59) | 0.52 (0.46 - 0.59) | 0.89 (0.74 - 1.07) | 0.93 (0.78 - 1.12) |
| Outpatient | 0.81 (0.75 - 0.89) | 0.82 (0.75 - 0.90) | 0.52 (0.45 - 0.59) | 0.52 (0.45 - 0.59) | 0.90 (0.74 - 1.08) | 0.93 (0.78 - 1.13) |
| Hospitalizations | 0.87 (0.69 - 1.12) | 0.86 (0.67 - 1.09) | 0.46 (0.33 - 0.66) | 0.46 (0.33 - 0.66) | 0.75 (0.37 - 1.55) | 0.83 (0.40 - 1.72) |

NA, not assessable

### Sensitivity analysis: lab-confirmed COVID-19

Analysis of lab-confirmed COVID-19 was performed as a sensitivity analysis, as rates of testing decreased substantially during the latter months of the study period. Data for the primary series mirrored that seen for the other outcomes, with mRNA-1273 showing higher point estimates for effectiveness than either BNT162b2 or Ad26.CoV.2 (rVE 28% and 52%, respectively; **Supplementary Table 6**). Subset analysis suggested no clear differences by age or region. Analysis of the booster dose was confounded by low rates of testing (see **Supplementary Table 6**) but also suggested increased effectiveness of mRNA-1273 versus BNT162b2.

## Discussion

To our knowledge, this is the first analysis of the rVE of mRNA-1273 compared with other COVID-19 vaccines which follows the same cohort of individuals through primary and booster vaccination. In order to emulate real-life decisions as much as possible, cohort entry dates into the booster phase of the study were based on recommended CDC dates and therefore analysis of the primary series was truncated from this point forward. Over a period where Delta predominated, a primary series of mRNA-1273 was more effective than either BNT162b2 or Ad26.COV2.S in preventing any medically-attended, outpatient, hospitalized, and lab-confirmed COVID-19. In addition, evaluation of the impact of a homologous booster during an Omicron dominant period demonstrated greater effectiveness with mRNA-1273 versus BNT162b2 against any medically-attended, outpatient, hospitalized, and lab-confirmed COVID-19. These differences appeared consistent by region and age, however, an mRNA-1273 booster appeared to have increased rVE against medically-attended COVID-19 compared with BNT162b2 in a subgroup analysis of older adults (≥65 years of age).

The results following the primary series are consistent with findings from previous real-world evaluations of rVE of COVID-19 vaccines. In a study of health records of US veterans who received a primary series of COVID-19 vaccine, mRNA-1273 was more effective than BNT162b2 in adults <65 years and > 65 years against symptomatic infection (57.3% vs 22.5% and 36.2% vs -23.3%, respectively), hospitalization (83.1% vs 57.0% and 64.7% vs 1.7%, respectively), and ICU admission or death (84.4% vs 66.4% and 73.8% vs 29.3%, respectively).[16]. Consistent with these findings, the authors also demonstrated mRNA1273 to be more effective than BNT162b2 in veterans with >1 chronic disease. In addition, evaluation of COVID-19 hospitalizations across 21 US states between March and August 2021 showed significantly higher vaccine effectiveness of a primary series of mRNA-1273 (93%; 95% CI: 91–95%) compared with BNT162b2 (88%; 85–91%) or Ad26.COV2.S (71%; 56–81%) [24].

A number of real-world studies have also demonstrated a clear impact of booster doses on vaccine effectiveness, particularly against the Omicron variant [12, 13, 25-28]. In a comparative study of booster vaccinations based on data from the OpenSAFELY-TPP database in the UK during the period of Delta and Omicron dominance, HR for mRNA-1273 versus BNT162b2 were 0.92 (95% CI: 0.91–0.92) and 0.67 (0.58–0.78) for lab-confirmed and hospitalized COVID-19, respectively, 12 weeks after booster vaccination [18]. Similarly, in a study in veterans in the US, 16-week risk of COVID-19-related outcomes was lower in recipients of a third dose of mRNA-1273 vs BNT162b2, particularly for documented infection (excess events for BNT162b2 vs mRNA-1273 was 45.5 (95% CI: 19.4–84.7) per 10,000 persons) [27].

In our study, exploratory analysis suggested an increased benefit of mRNA-1273 versus BNT162b2 booster vaccination against medically-attended COVID-19 in older adults (≥65 years). These differences in effectiveness would have a meaningful impact on preventing and reducing the burden of COVID-19 in individuals at higher risk of more severe disease. Consistent with these findings, an analysis by Mayr et al [16] demonstrated mRNA-1273 to be more effective than BNT162b2 in older veterans and those with chronic diseases. Evaluation of long-term antibody persistence following primary series and homologous boosting with mRNA COVID-19 vaccines has shown statistically higher antibody titers and persistence for mRNA-1273 compared with BNT162b2 against both the ancestral strain and subsequent variants [29]. While not assessed specifically in older adults, the higher titers and increased persistence may also contribute to greater effectiveness, which is particularly important in combination with immunosenescence in the older age group. While there was no significant difference in vaccine effectiveness between the mRNA vaccine boosters in younger adults (18–64 years), point estimates of HRs favored mRNA-1273 and may have been significant with larger sample sizes, as the majority of booster recipients prioritized during the period of this analysis were in the older age group (≥50 years). Future analysis including more recipients from the younger age groups will help to confirm this finding, and any differences in vaccine effectiveness within groups at high risk for severe disease.

A key strength of this analysis was the richness of the available data from EHRs, which include demographics, comorbidities, laboratory results, and healthcare encounters in both outpatient and hospital settings. This allowed close matching of individuals across multiple potential confounding variables. Additionally, the large number of individuals included in the analysis allowed robust subgroup assessments by age group and region. However, it should be noted that some age groups still had small sample sizes or low numbers of cases (e.g. lab-confirmed COVID-19 in patients ≥75 years) or lab tests in the booster phase. The results of this analysis should be interpreted within the context of the retrospective nature of the study. In the absence of randomization, there may be unmeasured differences between groups which may have confounded vaccine effectiveness estimates. An additional limitation of this type of study is that misclassification of exposure and outcomes are potentially more common than in a randomized clinical trial, although misclassification of the vaccine administration was unlikely because of comprehensive recording of the vaccine administration in our database and the strict time window for administration of the second dose. Furthermore, while clinical cases were determined in this analysis from EHR records, rather than directly identified following a positive PCR test, these were confirmed by a record of a positive PCR test in >90% of cases. Finally, there is a potential bias due to right-censoring of the data. As individuals included received vaccinations up until the end date, the follow-up period for some individuals is potentially very short, meaning that some medically-attended cases that occurred outside of the follow-up window are not included in this analysis.

In summary, receipt of a primary series of mRNA-1273 vaccine resulted in lower risk of any medically-attended, outpatient, hospitalized, and lab-confirmed COVID-19 compared with BNT162b2 or Ad26.CoV2.S during a Delta-dominant period. Boosting with mRNA-1273 also resulted in reduced risk of any medically-attended, outpatient, and hospitalized COVID-19 compared with BNT162b2 during an Omicron-dominant period.

## Data Availability

Individual-level data reported in this study are not publicly shared. Upon request, and subject to review, Veradigm may provide the de-identified aggregate-level data that support the findings of this study. Deidentified data (including participant data as applicable) may be shared upon approval of an analysis proposal and a signed data access agreement. Individual-level data reported in this study are not shared publicly but they are shared fully with regulatory agencies.

## Acknowledgements

The authors would like to thank the following individuals for their contribution: Editorial assistance was provided by Dr Jenny Engelmoer (Sula Communications BV)

