## Supplementary Information for "Relative effectiveness of BNT162b2, mRNA-1273, and Ad26.COV2.S vaccines and homologous boosting in preventing COVID-19 in adults in the US"

**Supplementary Appendix**

**Additional methodology on patient-level de-identification**

Patient-level de-identification was performed using an algorithm developed by Datavant (San Francisco, CA, USA), with tokens created separately for each dataset using fields including name, date of birth, and gender. Each data asset meets the minimum protected health information (PHI) data requirements as specified for use by the algorithm. Research staff were not involved in the preparation of datasets containing PHI or the running of the algorithm. The final linked data set was a merge of the patient-level de-identified tokens in each individual dataset and contained no PHI. This linked, de-identified dataset was compliant with the Health Insurance Portability and Accountability Act (HIPAA) and statistically certified for research use.

**Supplementary Table 1.** CVX, CPT, and NDC codes used to identify COVID-19 vaccines in the dataset

| **COVID-19 vaccine type** | **CPT** | **CVX** | **NDC** |
| --- | --- | --- | --- |
| **mRNA-1273** | 91301-0011A, 91301-0012A | 207 | 80777027310, 80777027315, 80777027398, 80777027399, 8077727315, 8077727398, 8077727399, 8077727310 |
| **BNT162-B2** | 91300-0001A, 91300-0002A  [91305](https://www.ama-assn.org/find-covid-19-vaccine-codes)  [91307](https://www.ama-assn.org/find-covid-19-vaccine-codes) | 208, 217, 218, 219 | 59267100001, 59267100002, 59267100003, 5926710001, 5926710002, 5926710003  59267-1055-159267-1055-01  59267-1025-01 |
| **Ad26.COV2.S** | 91303 | 212 | 59676-580-05  59676-580-15 |
| **3rd dose** |  |  |  |
| **mRNA-1273 (3rd dose)** | 91301 | 207 | 80777-273-10  80777-0273-10 |
| **BNT162-B2 (3rd dose)** | 91300  91305 |  | 59267-100-01  59267-1000-01 |
| **Booster** |  |  |  |
| **mRNA-1273 (booster)** | 91306 | 207 | 80777-273-10  80777-0273-10 |
| **BNT162-B2 (booster)** | 91305 | 208, 217, 218, 219 | 59267-1025-1  59267-1025-01 |
| **Ad26.COV2.S (booster)** | 91303 | 212 | 59676-580-05  59676-0580-05 |

**Supplementary Table 2.** ICD-10 and SNOMED codes used for identifying COVID-19-related medical encounters

|  | **Codes for COVID-related medical encounters** |
| --- | --- |
| **ICD-10-CM** | J1282, U071, U072, B34.2 |
| **SNOMED** | 1119302008, 119731000146105, 119741000146102, 119751000146104, 119981000146107, 1240411000000107, 1240521000000100, 1240531000000103, 1240541000000107, 1240561000000108, 1240581000000104, 674814021000119106, 840533007, 840534001, 840536004, 840539006, 866151004, 866152006, 870577009, 870588003, 870589006, 870590002, 870591003, 871562009 |

**Supplementary Table 3**. Baseline characteristics of matched patients included in the comparisons between the primary series of mRNA-1273 vs BNT162b2 and Ad26.COV2.S COVID-19 vaccines

|  |  | mRNA-1273 | BNT162b2 | SMD | mRNA-1273 | AD26.COV2.S | SMD |
| --- | --- | --- | --- | --- | --- | --- | --- |
| Number of patients |  | 1529930 | 1529930 |  | 484795 | 484795 |  |
| Age at index (mean (SD)) |  | 48.16 (15.71) | 48.08 (15.78) | 0.005 | 48.65 (15.44) | 48.6 (15.44) | 0.003 |
| Age group at index | 18-24 | 130240 (8.5) | 130240 (8.5) | 0 | 38789 (8.0) | 38789 (8.0) | 0 |
|  | 25-29 | 104378 (6.8) | 104378 (6.8) |  | 31223 (6.4) | 31223 (6.4) |  |
|  | 30-34 | 120128 (7.9) | 120128 (7.9) |  | 34744 (7.2) | 34744 (7.2) |  |
|  | 35-39 | 129245 (8.4) | 129245 (8.4) |  | 38696 (8.0) | 38696 (8.0) |  |
|  | 40-44 | 138849 (9.1) | 138849 (9.1) |  | 43271 (8.9) | 43271 (8.9) |  |
|  | 45-49 | 139692 (9.1) | 139692 (9.1) |  | 45535 (9.4) | 45535 (9.4) |  |
|  | 50-54 | 165594 (10.8) | 165594 (10.8) |  | 56802 (11.7) | 56802 (11.7) |  |
|  | 55-59 | 188592 (12.3) | 188592 (12.3) |  | 64901 (13.4) | 64901 (13.4) |  |
|  | 60-64 | 202330 (13.2) | 202330 (13.2) |  | 67233 (13.9) | 67233 (13.9) |  |
|  | 65-69 | 101887 (6.7) | 101887 (6.7) |  | 30258 (6.2) | 30258 (6.2) |  |
|  | 70-74 | 52953 (3.5) | 52953 (3.5) |  | 15631 (3.2) | 15631 (3.2) |  |
|  | 75-79 | 27830 (1.8) | 27830 (1.8) |  | 8420 (1.7) | 8420 (1.7) |  |
|  | 80-84 | 15433 (1) | 15433 (1) |  | 4759 (1.0) | 4759 (1.0) |  |
|  | 85-89 | 12779 (0.8) | 12779 (0.8) |  | 4533 (0.9) | 4533 (0.9) |  |
| Gender | Female | 872183 (57.0) | 872183 (57.0) | 0 | 253253 (52.2) | 253253 (52.2) | 0 |
|  | Male | 657747 (43.0) | 657747 (43.0) |  | 231542 (47.8) | 231542 (47.8) |  |
| Race | Black | 80946 (5.3) | 80946 (5.3) | 0 | 20494 (4.2) | 20494 (4.2) | 0 |
|  | White | 66639 (4.4) | 66639 (4.4) |  | 14700 (3.0) | 14700 (3.0) |  |
|  | Other | 594138 (38.8) | 594138 (38.8) |  | 200428 (41.3) | 200428 (41.3) |  |
|  | Unknown | 788207 (51.5) | 788207 (51.5) |  | 249173 (51.4) | 249173 (51.4) |  |
| Ethnicity | Hispanic | 86223 (5.6) | 90361 (5.9) | 0.012 | 27889 (5.8) | 27889 (5.8) | 0.059 |
|  | Non-Hispanic | 1212541 (79.3) | 1207461 (78.9) |  | 384079 (79.2) | 384079 (79.2) |  |
|  | Unknown | 231166 (15.1) | 232108 (15.2) |  | 72827 (15.0) | 72827 (15.0) |  |
| Insurance type | Commercial | 814772 (53.3) | 885506 (57.9) | 0.108 | 262766 (54.2) | 253436 (52.3) | 0.062 |
|  | Medicaid | 253693 (16.6) | 231867 (15.2) |  | 76783 (15.8) | 74192 (15.3) |  |
|  | Medicare Advantage | 116052 (7.6) | 100940 (6.6) |  | 35240 (7.3) | 35024 (7.2) |  |
|  | Medicare FFS | 8143 (0.5) | 6676 (0.4) |  | 2654 (0.5) | 2579 (0.5) |  |
|  | Other | 33020 (2.2) | 41125 (2.7) |  | 11654 (2.4) | 11918 (2.5) |  |
|  | Unknown | 304250 (19.9) | 263816 (17.2) |  | 95698 (19.7) | 107646 (22.2) |  |
| Region | Midwest | 332733 (21.7) | 332733 (21.7) | 0 | 98677 (20.4) | 98677 (20.4) | 0 |
|  | Northeast | 307677 (20.1) | 307677 (20.1) |  | 104420 (21.5) | 104420 (21.5) |  |
|  | South | 514228 (33.6) | 514228 (33.6) |  | 162570 (33.5) | 162570 (33.5) |  |
|  | West | 280045 (18.3) | 280045 (18.3) |  | 87265 (18.0) | 87265 (18.0) |  |
|  | Unknown | 95247 (6.2) | 95247 (6.2) |  | 31863 (6.6) | 31863 (6.6) |  |
| Community transmission level at index | High | 735733 (48.1) | 723981 (47.3) | 0.019 | 229164 (47.3) | 233571 (48.2) | 0.021 |
|  | Substantial | 393248 (25.7) | 399166 (26.1) |  | 127884 (26.4) | 124049 (25.6) |  |
|  | Moderate | 295948 (19.3) | 302668 (19.8) |  | 92735 (19.1) | 92424 (19.1) |  |
|  | Low | 9649 (0.6) | 8730 (0.6) |  | 3105 (0.6) | 2840 (0.6) |  |
|  | Not available | 95352 (6.2) | 95385 (6.2) |  | 31907 (6.6) | 31911 (6.6) |  |
| Month of index | 2-2021 | 1600 (0.1) | 3863 (0.3) | 0.037 | 325 (0.1) | 337 (0.1) | 0.003 |
|  | 3-2021 | 254706 (16.6) | 249197 (16.3) |  | 116404 (24) | 115895 (23.9) |  |
|  | 4-2021 | 564616 (36.9) | 567611 (37.1) |  | 158217 (32.6) | 158725 (32.7) |  |
|  | 5-2021 | 343175 (22.4) | 345304 (22.6) |  | 86664 (17.9) | 86680 (17.9) |  |
|  | 6-2021 | 135406 (8.9) | 133567 (8.7) |  | 43531 (9) | 43515 (9) |  |
|  | 7-2021 | 50640 (3.3) | 50601 (3.3) |  | 25799 (5.3) | 25788 (5.3) |  |
|  | 8-2021 | 70721 (4.6) | 70870 (4.6) |  | 27807 (5.7) | 27803 (5.7) |  |
|  | 9-2021 | 84119 (5.5) | 83864 (5.5) |  | 17488 (3.6) | 17517 (3.6) |  |
|  | 10-2021 | 24947 (1.6) | 25053 (1.6) |  | 8560 (1.8) | 8535 (1.8) |  |
| Number of OP visits (mean (SD)) |  | 1.54 (4.64) | 1.56 (4.63) | 0.005 | 1.54 (4.61) | 1.39 (4.30) | 0.035 |
| Number of hospitalizations (mean (SD)) |  | 0.11 (0.62) | 0.11 (0.66) | 0.003 | 0.11 (0.62) | 0.12 (0.70) | 0.017 |
| Number of distinct immunocompromising medications (mean (SD)) |  | 0.04 (0.22) | 0.04 (0.22) | 0.001 | 0.04 (0.23) | 0.03 (0.21) | 0.029 |
| Any comorbidity | No | 745938 (48.8) | 758960 (49.6) | 0.017 | 233688 (48.2) | 249516 (51.5) | 0.065 |
|  | Yes | 783992 (51.2) | 770970 (50.4) |  | 251107 (51.8) | 235279 (48.5) |  |
| Cancer | No | 1262258 (82.5) | 1247330 (81.5) | 0.025 | 398219 (82.1) | 404775 (83.5) | 0.036 |
|  | Yes | 267672 (17.5) | 282600 (18.5) |  | 86576 (17.9) | 80020 (16.5) |  |
| Cerebrovascular disease | No | 1489305 (97.3) | 1491173 (97.5) | 0.008 | 471331 (97.2) | 472098 (97.4) | 0.010 |
|  | Yes | 40625 (2.7) | 38757 (2.5) |  | 13464 (2.8) | 12697 (2.6) |  |
| Chronic kidney disease | No | 1478213 (96.6) | 1481254 (96.8) | 0.001 | 467749 (96.5) | 469279 (96.8) | 0.018 |
|  | Yes | 51717 (3.4) | 48676 (3.2) |  | 17046 (3.5) | 15516 (3.2) |  |
| Chronic lung disease | No | 1455348 (95.1) | 1465790 (95.8) | 0.033 | 460099 (94.9) | 460453 (95.0) | 0.003 |
|  | Yes | 74582 (4.9) | 64140 (4.2) |  | 24696 (5.1) | 24342 (5.0) |  |
| Liver disease | No | 1516988 (99.2) | 1517774 (99.2) | 0.006 | 480528 (99.1) | 480609 (99.1) | 0.002 |
|  | Yes | 12942 (0.8) | 12156 (0.8) |  | 4267 (0.9) | 4186 (0.9) |  |
| Diabetes mellitus (Type 1 or 2) | No | 1314811 (85.9) | 1331137 (87.0) | 0.031 | 413132 (85.2) | 422707 (87.2) | 0.057 |
|  | Yes | 215119 (14.1) | 198793 (13.0) |  | 71663 (14.8) | 62088 (12.8) |  |
| Heart disease | No | 1422383 (93.0) | 1428470 (93.4) | 0.016 | 448694 (92.6) | 451323 (93.1) | 0.021 |
|  | Yes | 107547 (7.0) | 101460 (6.6) |  | 36101 (7.4) | 33472 (6.9) |  |
| Psychiatric condition | No | 1264432 (82.6) | 1282529 (83.8) | 0.032 | 402156 (83.0) | 402301 (83.0) | 0.008 |
|  | Yes | 265498 (17.4) | 247401 (16.2) |  | 82639 (17.0) | 82494 (17.0) |  |
| Pregnancy | No | 1514163 (99.0) | 1512461 (98.9) | 0.011 | 480666 (99.1) | 481365 (99.3) | 0.016 |
|  | Yes | 15767 (1.0) | 17469 (1.1) |  | 4129 (0.9) | 3430 (0.7) |  |
| Any immunocompromised status | No | 1453488 (95.0) | 1453720 (95.0) | 0.001 | 459005 (94.7) | 462913 (95.5) | 0.037 |
|  | Yes | 76442 (5.0) | 76210 (5.0) |  | 25790 (5.3) | 21882 (4.5) |  |
| IC - General | No | 1455797 (95.2) | 1456126 (95.2) | 0.001 | 459800 (94.8) | 463611 (95.6) | 0.037 |
|  | Yes | 74133 (4.8) | 73804 (4.8) |  | 24995 (5.2) | 21184 (4.4) |  |
| IC - Blood transplant | No | 1525734 (99.7) | 1525477 (99.7) | 0.003 | 483313 (99.7) | 483689 (99.8) | 0.015 |
|  | Yes | 4196 (0.3) | 4453 (0.3) |  | 1482 (0.3) | 1106 (0.2) |  |
| IC - Organ transplant | No | 1523747 (99.6) | 1523488 (99.6) | 0.003 | 482558 (99.5) | 483237 (99.7) | 0.022 |
|  | Yes | 6183 (0.4) | 6442 (0.4) |  | 2237 (0.5) | 1558 (0.3) |  |
| IC - Malignancy | No | 1510753 (98.7) | 1509526 (98.7) | 0.007 | 478317 (98.7) | 479249 (98.9) | 0.017 |
|  | Yes | 19177 (1.3) | 20404 (1.3) |  | 6478 (1.3) | 5546 (1.1) |  |
| Current smoker | No | 1296085 (84.7) | 1324728 (86.6) | 0.053 | 409760 (84.5) | 402260 (83.0) | 0.042 |
|  | Yes | 233845 (15.3) | 205202 (13.4) |  | 75035 (15.5) | 82535 (17.0) |  |
| Former smoker | No | 1327555 (86.8) | 1331646 (87.0) | 0.008 | 419970 (86.6) | 421143 (86.9) | 0.007 |
|  | Yes | 202375 (13.2) | 198284 (13.0) |  | 64825 (13.4) | 63652 (13.1) |  |
| CCI score (mean (SD)) |  | 0.66 (1.33) | 0.63 (1.31) | 0.025 | 0.70 (1.37) | 0.63 (1.32) | 0.050 |
| CCI - Myocardial infarction | No | 1509075 (98.6) | 1510279 (98.7) | 0.007 | 477806 (98.6) | 477972 (98.6) | 0.003 |
|  | Yes | 20855 (1.4) | 19651 (1.3) |  | 6989 (1.4) | 6823 (1.4) |  |
| CCI - Congestive heart failure | No | 1487014 (97.2) | 1489746 (97.4) | 0.011 | 470487 (97.0) | 470953 (97.1) | 0.006 |
|  | Yes | 42916 (2.8) | 40184 (2.6) |  | 14308 (3.0) | 13842 (2.9) |  |
| CCI - Peripheral vascular disease | No | 1466435 (95.8) | 1470819 (96.1) | 0.015 | 463731 (95.7) | 464889 (95.9) | 0.012 |
|  | Yes | 63495 (4.2) | 59111 (3.9) |  | 21064 (4.3) | 19906 (4.1) |  |
| CCI - Cerebrovascular disease | No | 1486636 (97.2) | 1488442 (97.3) | 0.007 | 470517 (97.1) | 471374 (97.2) | 0.011 |
|  | Yes | 43294 (2.8) | 41488 (2.7) |  | 14278 (2.9) | 13421 (2.8) |  |
| CCI - Dementia | No | 1519466 (99.3) | 1519511 (99.3) | <0.001 | 481172 (99.3) | 480748 (99.2) | 0.010 |
|  | Yes | 10464 (0.7) | 10419 (0.7) |  | 3623 (0.7) | 4047 (0.8) |  |
| CCI - Chronic pulmonary disease | No | 1360174 (88.9) | 1373232 (89.8) | 0.028 | 429619 (88.6) | 432804 (89.3) | 0.021 |
|  | Yes | 169756 (11.1) | 156698 (10.2) |  | 55176 (11.4) | 51991 (10.7) |  |
| CCI - Rheumatic disease | No | 1494551 (97.7) | 1495325 (97.7) | 0.003 | 473357 (97.6) | 474330 (97.8) | 0.014 |
|  | Yes | 35379 (2.3) | 34605 (2.3) |  | 11438 (2.4) | 10465 (2.2) |  |
| CCI - Peptic ulcer disease | No | 1520036 (99.4) | 1520474 (99.4) | 0.004 | 481559 (99.3) | 481693 (99.4) | 0.003 |
|  | Yes | 9894 (0.6) | 9456 (0.6) |  | 3236 (0.7) | 3102 (0.6) |  |
| CCI - Mild liver disease | No | 1462461 (95.6) | 1464638 (95.7) | 0.007 | 462852 (95.5) | 464406 (95.8) | 0.016 |
|  | Yes | 67469 (4.4) | 65292 (4.3) |  | 21943 (4.5) | 20389 (4.2) |  |
| CCI - Diabetes without chronic complications | No | 1322782 (86.5) | 1338370 (87.5) | 0.030 | 415708 (85.7) | 425203 (87.7) | 0.056 |
|  | Yes | 207148 (13.5) | 191560 (12.5) |  | 69087 (14.3) | 59592 (12.3) |  |
| CCI - Mild to moderate renal disease | No | 1473535 (96.3) | 1476380 (96.5) | 0.010 | 466180 (96.2) | 467892 (96.5) | 0.019 |
|  | Yes | 56395 (3.7) | 53550 (3.5) |  | 18615 (3.8) | 16903 (3.5) |  |
| CCI - Diabetes with chronic complications | No | 1450596 (94.8) | 1457763 (95.3) | 0.022 | 458298 (94.5) | 461743 (95.2) | 0.032 |
|  | Yes | 79334 (5.2) | 72167 (4.7) |  | 26497 (5.5) | 23052 (4.8) |  |
| CCI - Hemiplegia or paraplegia | No | 1522813 (99.5) | 1523388 (99.6) | 0.006 | 482334 (99.5) | 482415 (99.5) | 0.002 |
|  | Yes | 7117 (0.5) | 6542 (0.4) |  | 2461 (0.5) | 2380 (0.5) |  |
| CCI - Any malignancy | No | 1475307 (96.4) | 1472691 (96.3) | 0.009 | 466533 (96.2) | 469124 (96.8) | 0.029 |
|  | Yes | 54623 (3.6) | 57239 (3.7) |  | 18262 (3.8) | 15671 (3.2) |  |
| CCI - Moderate to severe liver disease | No | 1526275 (99.8) | 1526432 (99.8) | 0.002 | 483540 (99.7) | 483606 (99.8) | 0.003 |
|  | Yes | 3655 (0.2) | 3498 (0.2) |  | 1255 (0.3) | 1189 (0.2) |  |
| CCI - Severe renal disease | No | 1522025 (99.5) | 1522579 (99.5) | 0.005 | 482043 (99.4) | 482417 (99.5) | 0.011 |
|  | Yes | 7905 (0.5) | 7351 (0.5) |  | 2752 (0.6) | 2378 (0.5) |  |
| CCI - Metastatic solid tumor | No | 1522738 (99.5) | 1522010 (99.5) | 0.007 | 482286 (99.5) | 482709 (99.6) | 0.013 |
|  | Yes | 7192 (0.5) | 7920 (0.5) |  | 2509 (0.5) | 2086 (0.4) |  |
| CCI - AIDS | No | 1529131 (99.9) | 1529347 (100.0) | 0.007 | 484513 (99.9) | 484566 (100.0) | 0.005 |
|  | Yes | 799 (0.1) | 583 (0.0) |  | 282 (0.1) | 229 (0.0) |  |
| CCI - HIV | No | 1523698 (99.6) | 1524579 (99.7) | 0.009 | 482620 (99.6) | 482972 (99.6) | 0.011 |
|  | Yes | 6232 (0.4) | 5351 (0.3) |  | 2175 (0.4) | 1823 (0.4) |  |
| EFI score* | <5% | 75866 (5.0) | 78179 (5.1) | 0.009 | 22265 (4.6) | 20886 (4.3) | 0.020 |
|  | 5% to <20% | 113317 (7.4) | 110935 (7.3) |  | 34196 (7.1) | 34781 (7.2) |  |
|  | 20%+ | 21699 (1.4) | 21768 (1.4) |  | 7140 (1.5) | 7934 (1.6) |  |
|  | Under 65 (not calculated) | 1319048 (86.2) | 1319048 (86.2) |  | 421194 (86.9) | 421194 (86.9) |  |

CCI, Charlson comorbidity index; EFI, electronic frailty index; EHR, electronic health record; IC, immunocompromised; IP, inpatient; IQR, interquartile range; OP, outpatient; SD, standard deviation; SMD, standardized mean difference

*EFI score was only calculated in patients ≥ 65 years of age.

**Supplementary Table 4**. Baseline characteristics of matched patients included in the comparison between the mRNA-1273 vs BNT162b2 booster doses

|  |  | mRNA-1273 | BNT162b2 | SMD |
| --- | --- | --- | --- | --- |
| Number of patients |  | 368100 | 368100 |  |
| Age at index (mean (SD)) |  | 50.21 (14.48) | 50.13 (14.55) | 0.006 |
| Age group at index | 18-24 | 20465 (5.6) | 20465 (5.6) | 0 |
|  | 25-29 | 18588 (5) | 18588 (5) |  |
|  | 30-34 | 24150 (6.6) | 24150 (6.6) |  |
|  | 35-39 | 29093 (7.9) | 29093 (7.9) |  |
|  | 40-44 | 32341 (8.8) | 32341 (8.8) |  |
|  | 45-49 | 33872 (9.2) | 33872 (9.2) |  |
|  | 50-54 | 43036 (11.7) | 43036 (11.7) |  |
|  | 55-59 | 54414 (14.8) | 54414 (14.8) |  |
|  | 60-64 | 66856 (18.2) | 66856 (18.2) |  |
|  | 65-69 | 23102 (6.3) | 23102 (6.3) |  |
|  | 70-74 | 10779 (2.9) | 10779 (2.9) |  |
|  | 75-79 | 5640 (1.5) | 5640 (1.5) |  |
|  | 80-84 | 3274 (0.9) | 3274 (0.9) |  |
|  | 85-89 | 2490 (0.7) | 2490 (0.7) |  |
| Gender | Female | 208574 (56.7) | 208574 (56.7) | 0 |
|  | Male | 159526 (43.3) | 159526 (43.3) |  |
| Race | Black | 15567 (4.2) | 15567 (4.2) | 0 |
|  | White | 18251 (5.0) | 18251 (5.0) |  |
|  | Other | 145102 (39.4) | 145102 (39.4) |  |
|  | Unknown | 189180 (51.4) | 189180 (51.4) |  |
| Ethnicity | Hispanic | 16703 (4.5) | 17903 (4.9) | 0.015 |
|  | Non-Hispanic | 293318 (79.7) | 292658 (79.5) |  |
|  | Unknown | 58079 (15.8) | 57539 (15.6) |  |
| Insurance type | Commercial | 229516 (62.4) | 243602 (66.2) | 0.131 |
|  | Medicaid | 36942 (10.0) | 39183 (10.6) |  |
|  | Medicare Advantage | 23486 (6.4) | 20836 (5.7) |  |
|  | Medicare FFS | 2236 (0.6) | 2590 (0.7) |  |
|  | Other | 7062 (1.9) | 9261 (2.5) |  |
|  | Unknown | 68858 (18.7) | 52628 (14.3) |  |
| Region | Midwest | 74615 (20.3) | 74615 (20.3) | 0 |
|  | Northeast | 79382 (21.6) | 79382 (21.6) |  |
|  | South | 120090 (32.6) | 120090 (32.6) |  |
|  | West | 71448 (19.4) | 71448 (19.4) |  |
|  | Unknown | 22565 (6.1) | 22565 (6.1) |  |
| Community transmission level at index | High | 4206 (1.1) | 4238 (1.2) | 0.035 |
|  | Substantial | 4349 (1.2) | 3100 (0.8) |  |
|  | Moderate | 399 (0.1) | 314 (0.1) |  |
|  | Low | 359146 (97.6) | 360448 (97.9) |  |
|  | Not available | 4206 (1.1) | 4238 (1.2) |  |
| Month of index | 10-2021 | 34694 (9.4) | 31885 (8.7) | 0.028 |
|  | 11-2021 | 133461 (36.3) | 133312 (36.2) |  |
|  | 12-2021 | 153080 (41.6) | 155854 (42.3) |  |
|  | 1-2022 | 46865 (12.7) | 47049 (12.8) |  |
| Number of OP visits (mean (SD)) |  | 1.06 (3.39) | 1.61 (4.65) | 0.135 |
| Number of hospitalizations (mean (SD)) |  | 0.08 (0.50) | 0.09 (0.60) | 0.015 |
| Number of distinct immunocompromising medications (mean (SD)) |  | 0.03 (0.20) | 0.03 (0.20) | 0.007 |
| Any comorbidity | No | 175043 (47.6) | 169491 (46.0) | 0.030 |
|  | Yes | 193057 (52.4) | 198609 (54.0) |  |
| Cancer | No | 288217 (78.3) | 283701 (77.1) | 0.030 |
|  | Yes | 79883 (21.7) | 84399 (22.9) |  |
| Cerebrovascular disease | No | 358920 (97.5) | 358409 (97.4) | 0.009 |
|  | Yes | 9180 (2.5) | 9691 (2.6) |  |
| Chronic kidney disease | No | 355665 (96.6) | 355515 (96.6) | 0.002 |
|  | Yes | 12435 (3.4) | 12585 (3.4) |  |
| Chronic lung disease | No | 352845 (95.9) | 353684 (96.1) | 0.012 |
|  | Yes | 15255 (4.1) | 14416 (3.9) |  |
| Liver disease | No | 365118 (99.2) | 365116 (99.2) | <0.001 |
|  | Yes | 2982 (0.8) | 2984 (0.8) |  |
| Diabetes mellitus (Type 1 or 2) | No | 316640 (86.0) | 318490 (86.5) | 0.015 |
|  | Yes | 51460 (14.0) | 49610 (13.5) |  |
| Heart disease | No | 342664 (93.1) | 343072 (93.2) | 0.004 |
|  | Yes | 25436 (6.9) | 25028 (6.8) |  |
| Psychiatric condition | No | 311182 (84.5) | 309806 (84.2) | 0.010 |
|  | Yes | 56918 (15.5) | 58294 (15.8) |  |
| Pregnancy | No | 366276 (99.5) | 366181 (99.5) | 0.004 |
|  | Yes | 1824 (0.5) | 1919 (0.5) |  |
| Any immunocompromised status | No | 349698 (95.0) | 348872 (94.8) | 0.010 |
|  | Yes | 18402 (5.0) | 19228 (5.2) |  |
| IC - General | No | 350326 (95.2) | 349578 (95.0) | 0.009 |
|  | Yes | 17774 (4.8) | 18522 (5.0) |  |
| IC - Blood transplant | No | 367120 (99.7) | 366923 (99.7) | 0.010 |
|  | Yes | 980 (0.3) | 1177 (0.3) |  |
| IC - Organ transplant | No | 366944 (99.7) | 366730 (99.6) | 0.010 |
|  | Yes | 1156 (0.3) | 1370 (0.4) |  |
| IC - Malignancy | No | 363184 (98.7) | 362701 (98.5) | 0.011 |
|  | Yes | 4916 (1.3) | 5399 (1.5) |  |
| Current smoker | No | 324865 (88.3) | 326261 (88.6) | 0.012 |
|  | Yes | 43235 (11.7) | 41839 (11.4) |  |
| Former smoker | No | 321663 (87.4) | 321166 (87.2) | 0.004 |
|  | Yes | 46437 (12.6) | 46934 (12.8) |  |
| CCI score (mean (SD)) |  | 0.63 (1.25) | 0.64 (1.27) | 0.005 |
| CCI - Myocardial infarction | No | 363510 (98.8) | 363443 (98.7) | 0.002 |
|  | Yes | 4590 (1.2) | 4657 (1.3) |  |
| CCI - Congestive heart failure | No | 359078 (97.5) | 359027 (97.5) | 0.001 |
|  | Yes | 9022 (2.5) | 9073 (2.5) |  |
| CCI - Peripheral vascular disease | No | 353658 (96.1) | 353528 (96.0) | 0.002 |
|  | Yes | 14442 (3.9) | 14572 (4.0) |  |
| CCI - Cerebrovascular disease | No | 358314 (97.3) | 357721 (97.2) | 0.010 |
|  | Yes | 9786 (2.7) | 10379 (2.8) |  |
| CCI - Dementia | No | 366284 (99.5) | 365960 (99.4) | 0.012 |
|  | Yes | 1816 (0.5) | 2140 (0.6) |  |
| CCI - Chronic pulmonary disease | No | 330367 (89.7) | 330839 (89.9) | 0.004 |
|  | Yes | 37733 (10.3) | 37261 (10.1) |  |
| CCI - Rheumatic disease | No | 360470 (97.9) | 360315 (97.9) | 0.003 |
|  | Yes | 7630 (2.1) | 7785 (2.1) |  |
| CCI - Peptic ulcer disease | No | 365831 (99.4) | 365729 (99.4) | 0.004 |
|  | Yes | 2269 (0.6) | 2371 (0.6) |  |
| CCI - Mild liver disease | No | 351499 (95.5) | 351058 (95.4) | 0.006 |
|  | Yes | 16601 (4.5) | 17042 (4.6) |  |
| CCI - Diabetes without chronic complications | No | 318488 (86.5) | 320143 (87) | 0.013 |
|  | Yes | 49612 (13.5) | 47957 (13) |  |
| CCI - Mild to moderate renal disease | No | 355007 (96.4) | 354751 (96.4) | 0.004 |
|  | Yes | 13093 (3.6) | 13349 (3.6) |  |
| CCI - Diabetes with chronic complications | No | 349944 (95.1) | 350388 (95.2) | 0.006 |
|  | Yes | 18156 (4.9) | 17712 (4.8) |  |
| CCI - Hemiplegia or paraplegia | No | 366771 (99.6) | 366619 (99.6) | 0.007 |
|  | Yes | 1329 (0.4) | 1481 (0.4) |  |
| CCI - Any malignancy | No | 354391 (96.3) | 353311 (96.0) | 0.015 |
|  | Yes | 13709 (3.7) | 14789 (4.0) |  |
| CCI - Moderate to severe liver disease | No | 367322 (99.8) | 367289 (99.8) | 0.002 |
|  | Yes | 778 (0.2) | 811 (0.2) |  |
| CCI - Severe renal disease | No | 366849 (99.7) | 366827 (99.7) | 0.001 |
|  | Yes | 1251 (0.3) | 1273 (0.3) |  |
| CCI - Metastatic solid tumor | No | 366586 (99.6) | 366436 (99.5) | 0.006 |
|  | Yes | 1514 (0.4) | 1664 (0.5) |  |
| CCI - AIDS | No | 367974 (100.0) | 367966 (100.0) | 0.001 |
|  | Yes | 126 (0.0) | 134 (0.0) |  |
| CCI - HIV | No | 366953 (99.7) | 366974 (99.7) | 0.001 |
|  | Yes | 1147 (0.3) | 1126 (0.3) |  |
| EFI score* | <5% | 17387 (4.7) | 17829 (4.8) | 0.019 |
|  | 5% to <20% | 24006 (6.5) | 22989 (6.2) |  |
|  | 20%+ | 3892 (1.1) | 4467 (1.2) |  |
|  | Under 65 (not calculated) | 322815 (87.7) | 322815 (87.7) |  |

CCI, Charlson comorbidity index; EFI, electronic frailty index; EHR, electronic health record; IC, immunocompromised; IP, inpatient; IQR, interquartile range; OP, outpatient; SD, standard deviation; SMD, standardized mean difference

*EFI score was only calculated in patients ≥ 65 years of age.

**Supplementary Table 5.** Number (%) matched pairs per age category for each vaccine comparison after the primary series and booster dose

|  | **Primary series** | | **Booster** |
| --- | --- | --- | --- |
| **Age group** | **mRNA-1273 vs BNT162b2** | **mRNA-1273 vs Ad26.COV2.S** | **mRNA-1273 vs BNT162b2** |
| 18–64 years | 1,319,048 (75.8) | 421,194 (76.8) | 322,815 (87.7) |
| ≥65 years | 210,882 (12.1) | 63,601 (11.6) | 45,285 (12.3) |
| 65–74 years | 154,840 (8.9) | 45,889 (8.4) | 33,881 (9.2) |
| ≥75 years | 56,042 (3.2) | 17,712 (3.2) | 11,404 (3.1) |

**Supplementary Table 6**. Unadjusted and adjusted hazard ratios for mRNA-1273 with BNT162b2 and Ad26.COV2.S (primary series) and with BNT162b2 (booster dose) against lab-confirmed COVID-19 by age and region

|  | **Primary Series** | | | | **Booster** | |
| --- | --- | --- | --- | --- | --- | --- |
|  | **mRNA-1273 vs BNT162b2^a^** | | **mRNA-1273 vs Ad26.COV2.S^b^** | | **mRNA-1273 vs BNT162b2^c^** | |
| **Variable** | **Unadjusted HR** | **Adjusted HR** | **Unadjusted HR** | **Adjusted HR** | **Unadjusted HR** | **Adjusted HR** |
| Overall | 0.74 (0.68 - 0.81) | 0.72 (0.66 - 0.78) | 0.48 (0.42 - 0.55) | 0.48 (0.42 - 0.55) | 0.36 (0.28 – 0.45) | 0.39 (0.31 – 0.49) |
| Age group |  |  |  |  |  |  |
| 18-64 years | 0.78 (0.71 - 0.86) | 0.76 (0.69 - 0.83) | 0.45 (0.39 - 0.53) | 0.45 (0.39 - 0.53) | 0.39 (0.30 – 0.50) | 0.38 (0.29 – 0.49) |
| ≥65 years | 0.63 (0.53 - 0.75) | 0.60 (0.50 - 0.72) | 0.66 (0.48 - 0.92) | 0.66 (0.48 - 0.92) | 0.23 (0.13 – 0.43) | 0.31 (0.16 - 0.58) |
| 65-74 years | 0.59 (0.47 - 0.73) | 0.56 (0.45 - 0.69) | 0.67 (0.46 - 0.98) | 0.67 (0.46 - 0.98) | 0.23 (0.11 – 0.48) | 0.31 (0.15 – 0.65) |
| ≥75 years | 0.74 (0.54 - 1.02) | 0.72 (0.52 - 0.99) | 0.63 (0.33 - 1.23) | 0.63 (0.33 - 1.23) | NA (NA - NA) | NA (NA - NA) |
| Region |  |  |  |  |  |  |
| Midwest | 0.71 (0.60 - 0.84) | 0.71 (0.60 - 0.84) | 0.34 (0.26 - 0.46) | 0.34 (0.26 - 0.46) | 0.26 (0.15 - 0.44) | 0.31 (0.18 - 0.53) |
| Northeast | 0.92 (0.78 – 1.10) | 0.92 (0.79 - 1.10) | 0.63 (0.48 - 0.83) | 0.66 (0.50 - 0.87) | 0.45 (0.29 - 0.69) | 0.51 (0.33 - 0.78) |
| South | 0.71 (0.62 - 0.82) | 0.66 (0.57 - 0.75) | 0.57 (0.46 - 0.71) | 0.57 (0.46 - 0.71) | 0.30 (0.19 - 0.45) | 0.33 (0.22 - 0.51) |
| West | 0.58 (0.43 - 0.78) | 0.56 (0.42 - 0.76) | 0.27 (0.17 - 0.44) | 0.26 (0.16 - 0.43) | 0.52 (0.29 - 0.94) | 0.65 (0.36 - 1.19) |
| Unknown | 0.87 (0.49 - 1.54) | 0.85 (0.48 - 1.51) | 0.63 (0.30 - 1.30) | 0.63 (0.30 - 1.30) | NA (NA - NA ) | NA (NA - NA) |

^a^Number of lab-confirmed COVID-19 cases (PCR or antigen tests): mRNA-1273: 288,950; BNT162b2: 308,271

^b^Number of lab-confirmed COVID-19 cases (PCR or antigen tests): mRNA-1273: 91,698; Ad26.COV2.S: 95,293

^c^Number of lab-confirmed COVID-19 cases (PCR or antigen tests): mRNA-1273: 61,324; BNT162b2: 66,221

Lab-confirmed COVID-19 was defined as a lab test for COVID indicating a positive result as recorded in the EHR

**Supplementary Figure 1**. Comparison of the cumulative incidence of (A) overall COVID-19 (primary outcome), (B) outpatient COVID-19, (C) hospitalized COVID-19, and (D) lab-confirmed COVID-19 for mRNA-1273 vs BNT162b2 and Ad26.COV2.S

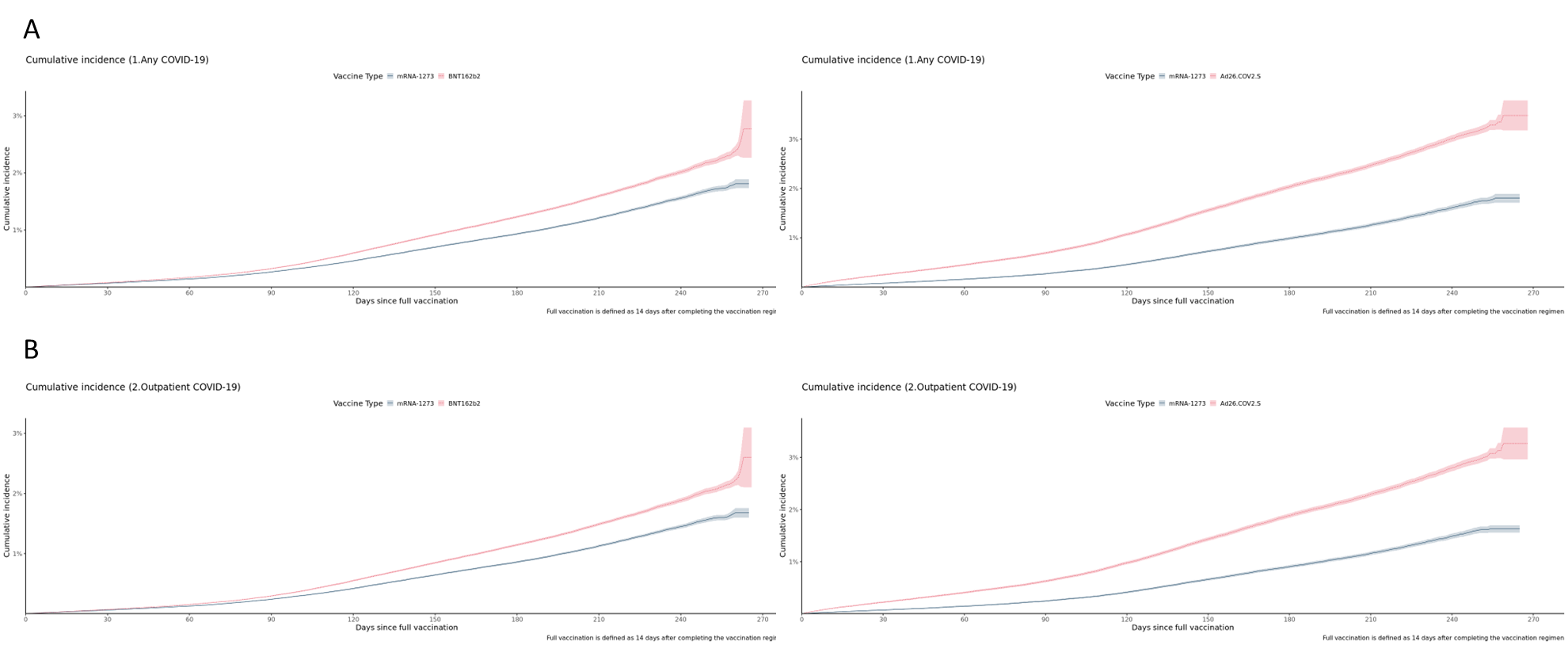

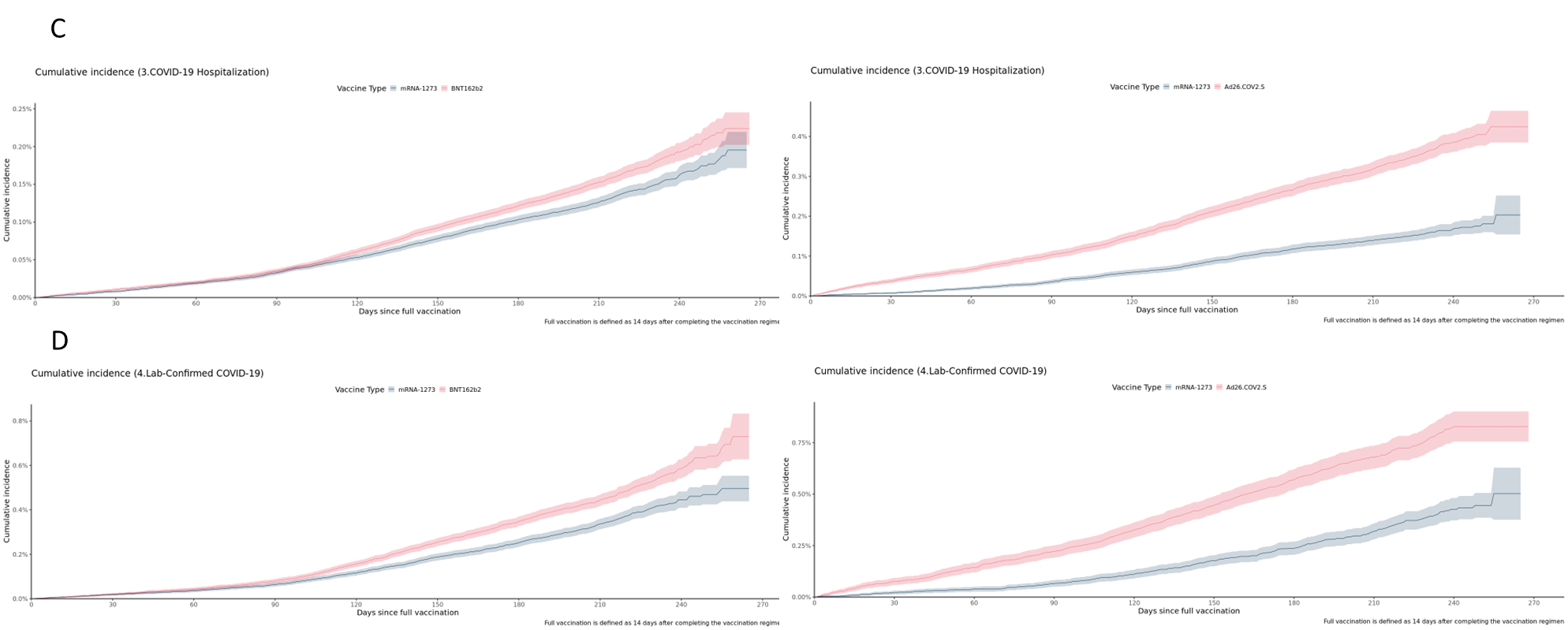

**Supplementary Figure 2**. Comparison of the cumulative incidence of (A) overall COVID-19 (primary outcome), (B) outpatient COVID-19, (C) hospitalized COVID-19, and (D) lab-confirmed COVID-19 for mRNA-1273 vs BNT162b2 booster dose

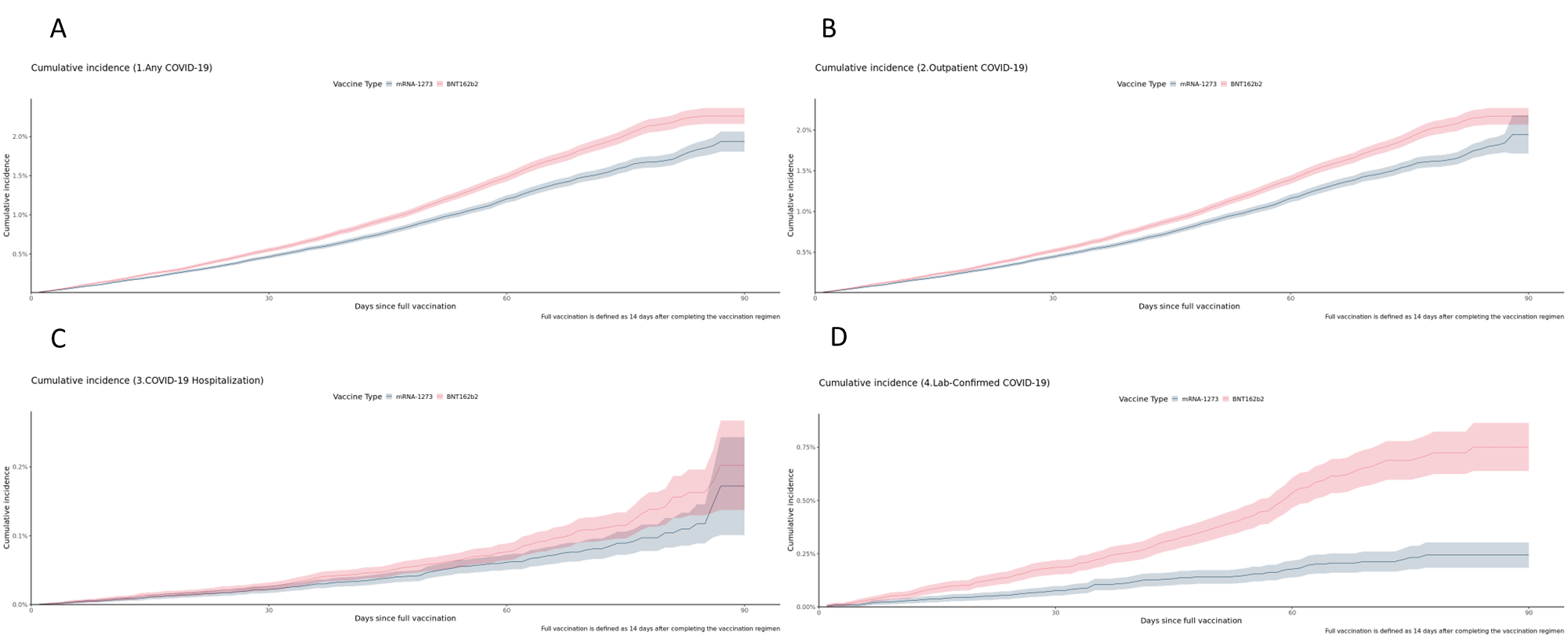
